# FAMILY LIFE EXPERIENCES IN CARING FOR CHILDREN WITH LEUKEMIA: A LITERATURE REVIEW

**DOI:** 10.1101/2022.01.18.22269327

**Authors:** Siti Nurjanah, Yurike Septianingrum, Moses Glorino Rumambo Pandin

**Affiliations:** Doctoral of Nursing, Faculty of Nursing, Universitas Airlangga, Jalan Dr. Ir. H. Soekarno, Mulyorejo, Kec. Mulyorejo, Surabaya, East Java 60115; Nursing Department, Faculty of Nursing, Universitas Nahdlatul Ulama Surabaya, Jalan SMEA No. 57 Surabaya, East Java 60237; Nursing Department, Faculty of Nursing, Universitas Airlangga, Jalan Dr. Ir. H. Soekarno, Mulyorejo, Kec. Mulyorejo, Surabaya, East Java 60115

**Keywords:** acute lymphoblastic leukemia, cancer, children, parent experiences

## Abstract

Acute Lymphoblastic Leukemia (ALL) is the most common childhood cancer, and ALL is the leading cause of death in children. Chronic diseases, one of which is cancer suffered by children, can provide varied responses to families. Caring for children with cancer requires a very long and complex process, so it requires good coordination between children, parents, families and the health team. The purpose of this literature review is to obtain in-depth information on the experiences of parents in caring for children with cancer. The method of writing this article is a literature review of 40 articles with the year 2019-2021 published from electronic database, namely CINAHL, Web of Science, SAGE and Proquest. The method of searching and selecting articles used The Center for Review and Dissemination and the Joanna Briggs Institute Guideline and PRISMA diagram. Checklist with selection criteria using the PICOS approach. The results of a literature review show that while caring for children with cancer, parents experience stress, anxiety about losing a child, uncertainty in child treatment, difficulties in caring for children and family responsibilities. Support in the form of psychosocial, material, and social support is needed for parents. The declining health condition of children is the biggest obstacle for parents, material needs are needed to support the care of children suffering from cancer, and there is hope from parents for their child’s recovery.

## INTRODUCTION

Currently, cancer is a serious disease and threatens the health of children in the world. According to data from the World Health Organization (WHO), every year there are 6.25 million people with cancer in the world, of which 4% or 250 thousand are children. Cancer is a disease that can be suffered by anyone, regardless of age, whether children, adolescents or adults. The incidence of cancer in children in Indonesia has also increased, namely 100,000 children. Child cancer data in Jakarta from 19,000 cases to 14 million with a death rate of 8.2 million cases (Nahata et al., 2019).

Chronic disease in this case cancer suffered by children can give different responses to families and this is influenced by experience. The impact of cancer suffered by children can cause parents to experience a psychological response which is very important to study because it can have an influence on other family members and the psychology of the child himself. The values held by the family and the ethnic or cultural background that comes from the ancestors will affect the belief in a disease. Each case has a different problem, this can be influenced by cultural, religious and ethnic backgrounds as well as the health management system that is not the same in every family (Lambert et al., 2021).

Every family with or without children with leukemia has problems that usually arise in the family, including financial problems, competition for attention between siblings, attention to children or other family members, and the ability to cope with important periods in the child’s development. Families who have children with leukemia will cause a heavy burden for other family members. Parents will feel guilty and feel responsible for what happened to their children, or even parents hope to replace and bear the disease suffered by their children. Parents need help and support from all parties involved, both for the welfare of the parents themselves and for efforts to heal and care for their children. Some parents will feel stressed and there are many things that must be known in recognizing and understanding the experiences of parents caring for children with leukemia (Bouffet et al., 2020).

Psychological discomfort that parents often feel as caregivers of children suffering from blood cancer or leukemia are feelings of anxiety and depression by parents (56%), guilt, fear, worry, sadness, depression will be felt for approximately 5 years and can be back to normal after a few years. The experience of parents in caring for children with cancer can be explored through qualitative studies. The qualitative method used in nursing is an approach that aims to obtain complete and specific, in-depth information and understand what parents are experiencing so that they can help and provide the support needed by patients and their families (SINAGA, 2017). Several factors contributed to treatment failure, namely economic factors, education level. Stress experienced by parents, lack of knowledge about side effects of treatment and how to deal with them and lack of experience in caring for children can also have an impact on parents’ ability to care for children with cancer. Therefore, information related to the experience of caring for and the need for information based on the perceptions of parents is very important in the success of the goal of successful treatment and care for children with cancer (Erker et al., 2018).

## METHOD

### Eligibillity criteria

Determine the criteria with an article search strategy with the PICOS framework (Population, Intervention, Comparison, Outcome, Study type) which is also adjusted to the inclusion and exclusion criteria (table 1) below

**Table 1.** Inclusion and Exclusion criteria in PICOS

| Criteria | Inclusion | Exclusion |
| --- | --- | --- |
| Population | Child (age 1-17 years) | Teenager, Adult |
| Intervention | Child nursing care | There is no criteria exclusion |
| Comparison | There is no comparison | There is no criteria exclusion |
| Outcome | Family experiences in caring of children with leukemia | No family experiences in caring of children with leukemia |
| Study type | Qualitative study | Cross sectional study, mixed method, quasi experimental design, clinical trial or randomized controlled trial, Systematic or literature reviews |

### Search strategy

The literature search begins by determining the keywords to be used using Boolean operators (AND, OR NOT or AND NOT). The keywords in this literature review were adjusted according to the Medical Subject Heading (MeSH) namely “leukemia” OR “acute lymphoblastic leukemia” AND “child” OR “children” AND “experience” OR “family experience” OR “parent experience”. The databases used in the literature search were Scopus, CINAHL, Web of science, SAGE and Proquest, namely English articles for the last 5 years, 2016 – 2021, obtained 170 records, then filtered to remove duplicates, 153 records were obtained. The screening of records was determined by the eligibility criteria with the article search strategy with the PICOS framework which was also adjusted to the inclusion and exclusion criteria (table 1), obtained 81 records. A total of 14 full-text articles were assessed for eligibility, and only 8 articles matched eligibility in the qualitative study.

**Figure 1.**
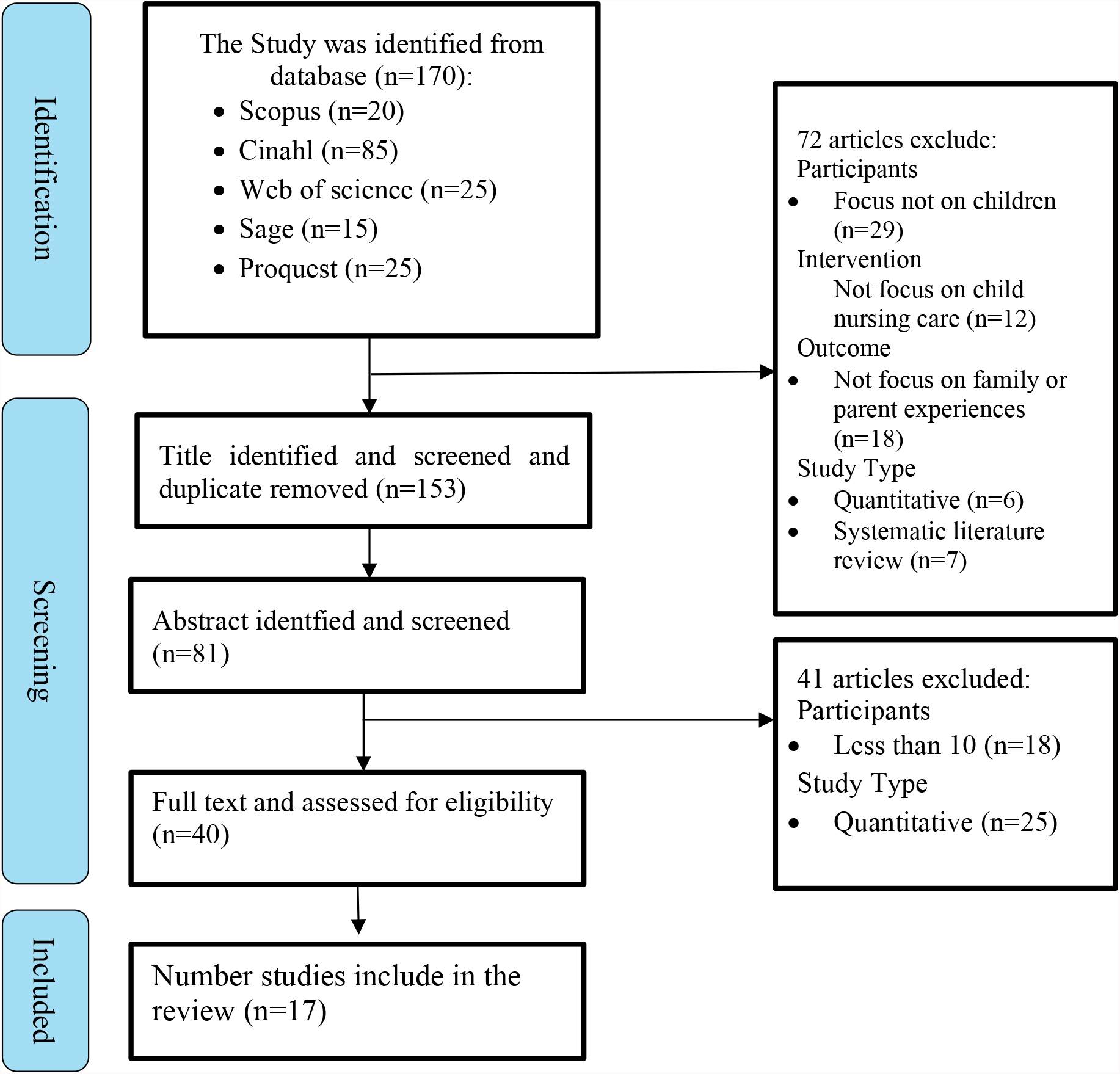
PRISMA Diagram of the Literature Search and Screening Process

## RESULT

**Table 2.**
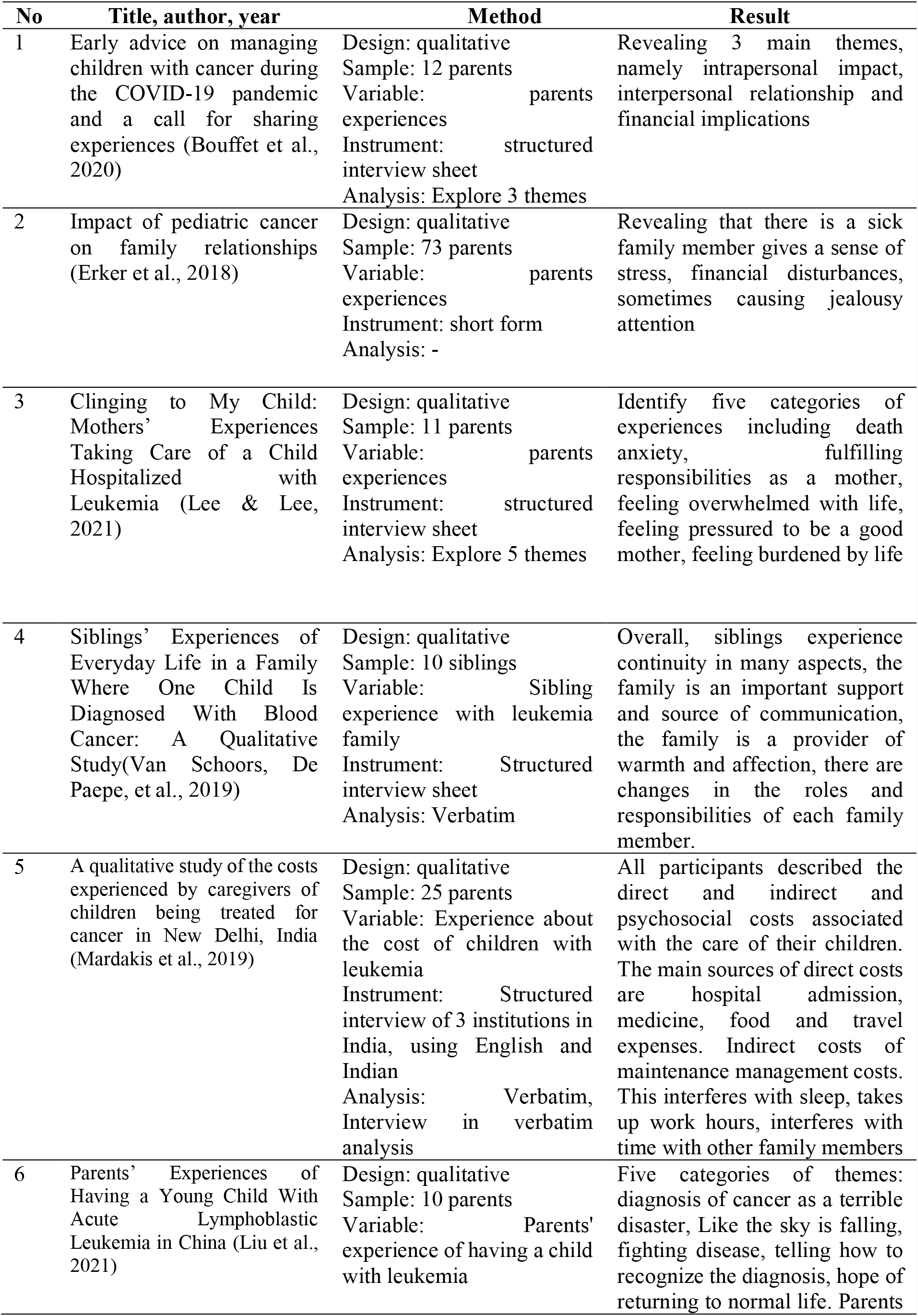
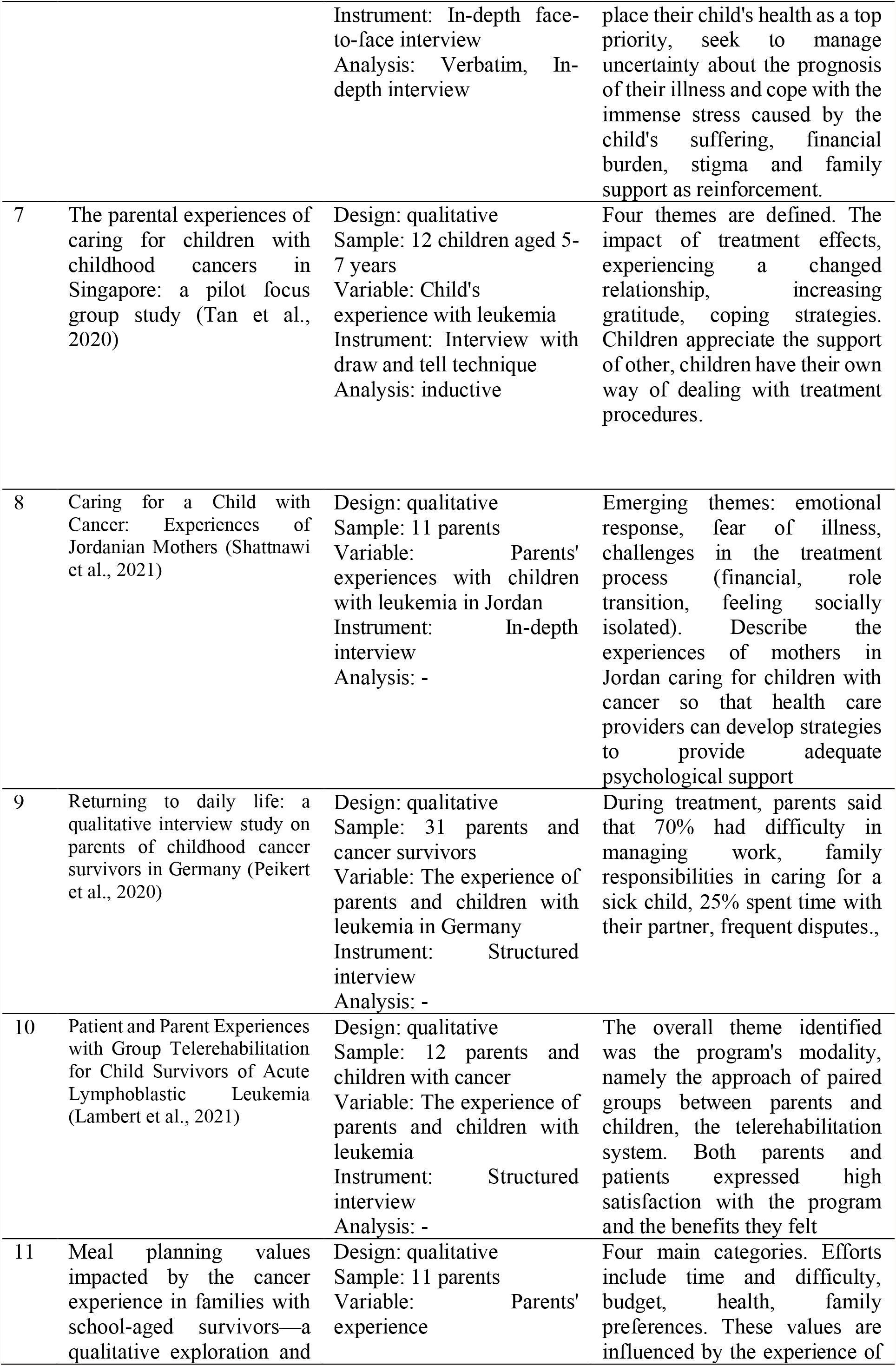
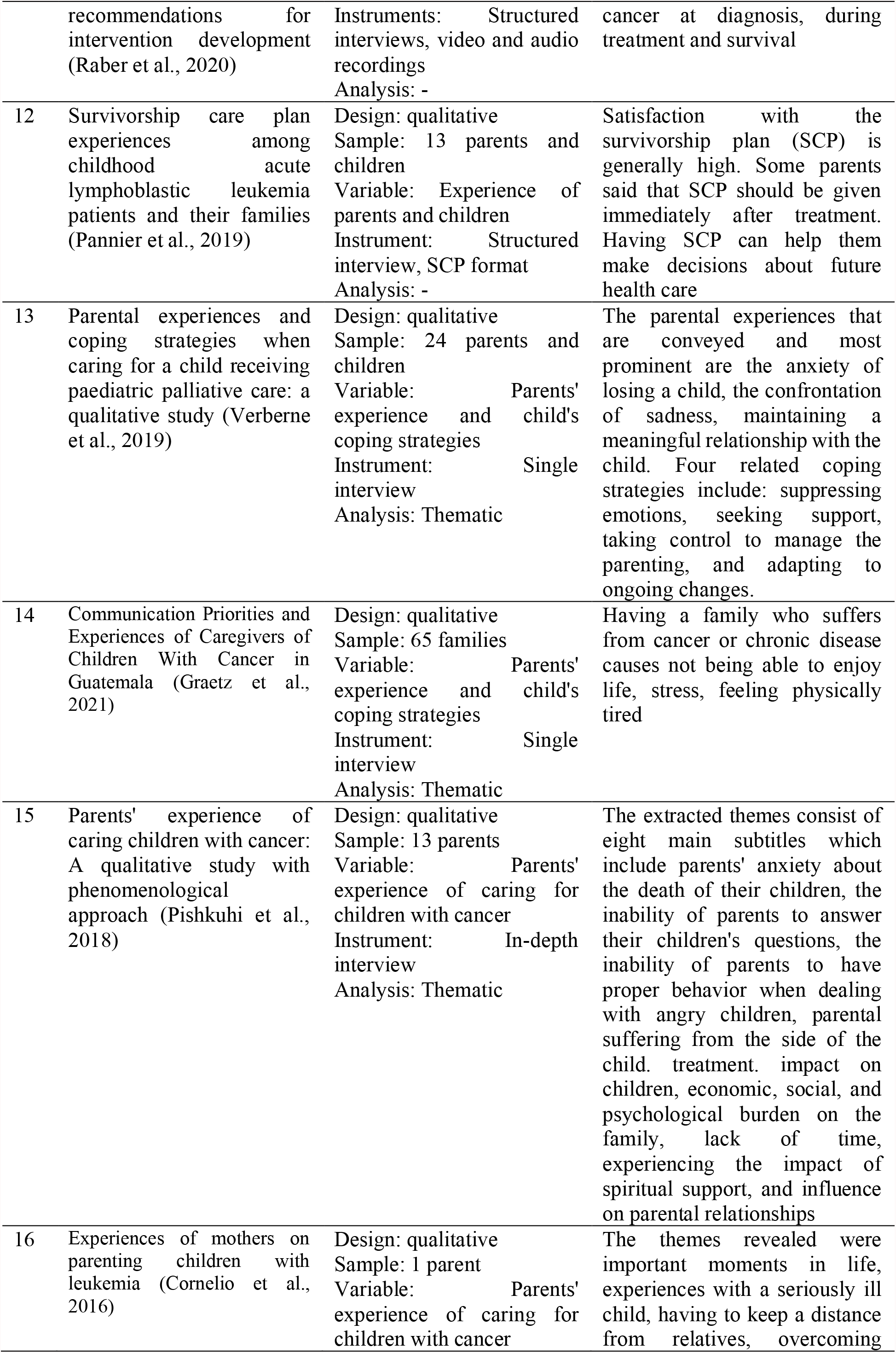
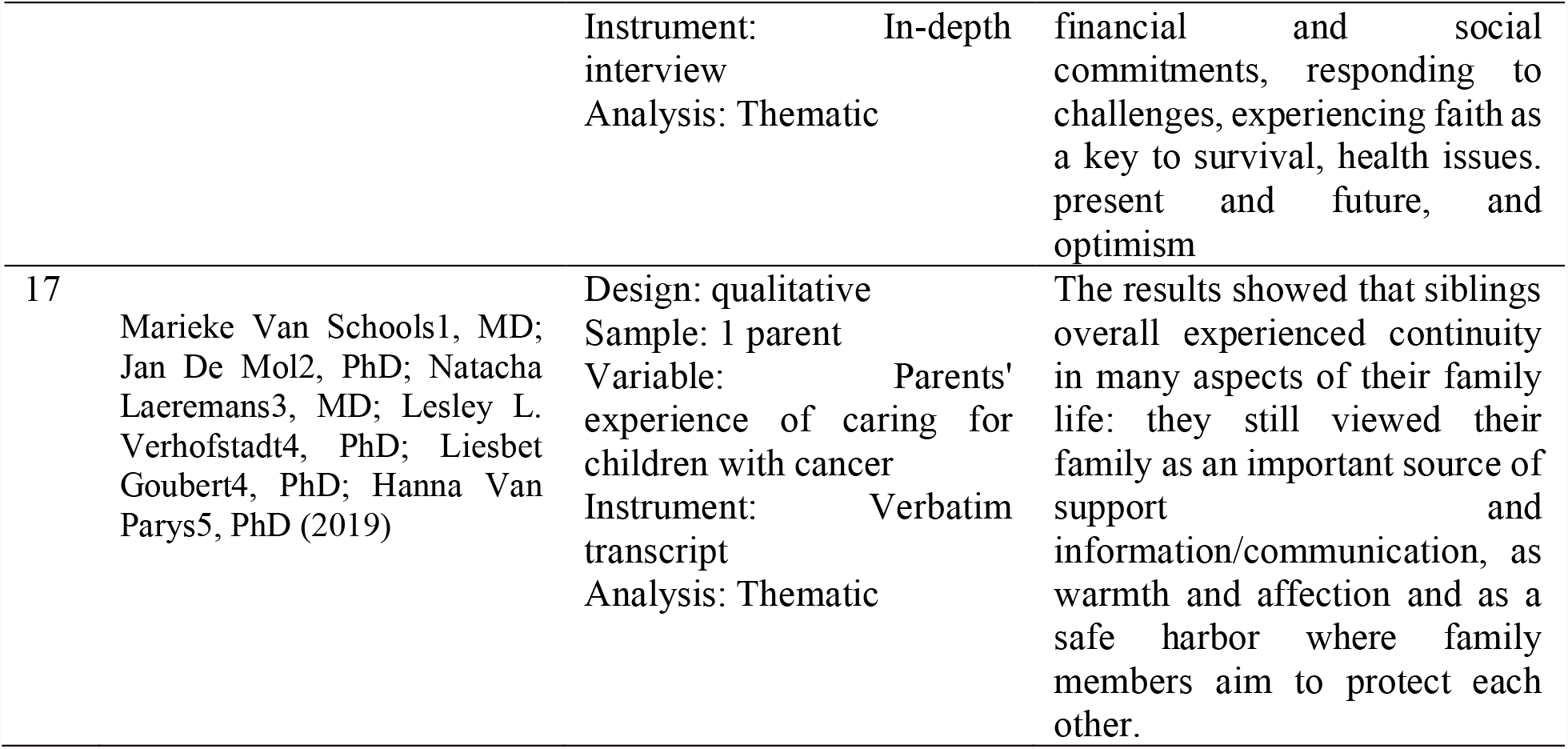
Literature search results

## DISCUSSION

The results of research that explores the experiences of parents, children’s experiences and family experiences in caring for children with cancer or leukemia convey that there is a sense of stress experienced by families, feelings of sadness, panic, disappointment and confusion over their child’s condition (Lambert et al., 2021).

In addition, there were four themes that were obtained, namely emotional responses: 1) “I feel sad today to hear that my child is sick with leukemia, I don’t know how I feel” (P1). “Yes, sadly, I can’t believe it (Craig E, Ke Yan, Liyun Z, Kristin B, Katryn E, 2020). Because they don’t feel any symptoms” (P2). “Yes, I feel sad (bowed down), how come my child is sick like this” (P3). 2) Attitude during treatment: “just go through the treatment process, following the treatment schedule that has been planned” (P1). “We have to put our trust and try to follow the treatment process, every day we have to drink juice and watch our food” (P2). “For now, it’s starting to feel a bit relieved, my child has started to feel less pain in the joints and is not vomiting anymore” (P3). 3) Support obtained: “There is support from my family, that’s what makes me enthusiastic and should not give up” (P1). “What kind of support is it, because I was only told to be patient, it’s good to say it because I didn’t experience it myself” (P2). Yes, there must be support, especially from my family and from people who share the same fate as me. And that is very helpful” (P3). 4) Illness Impact: “This is the first child, because we are sick like this, we have to postpone having a younger sibling. I’m afraid it won’t be taken care of later” (P1). “His father went to prison, brother, because we are economically pressed to pay for the treatment process and other needs so we look for shortcuts to do evil” (P2). “Yes, the impact on me. Because I have no appetite and can’t go anywhere. Every time there is a sense of anxiety and fear” (Chivukula et al., 2018).

Families, especially mothers, will face challenges in accepting and adjusting to children suffering from leukemia. In addition to adapting to the child’s condition, the mother and family also struggle to be able to deal with pressure during the treatment process, the high costs and confusion in dealing with their child’s future. So it takes support from various parties to provide motivation and psychological support for families with children with leukemia (Kyololo et al., 2019).

According to other research results through interviews to identify the experience of parents in caring for children with leukemia, there are eight main themes, namely 1) parental concerns about the child’s death, 2) the inability of parents to answer children’s questions, 3) the inability of parents to handle child aggression, 4) parental discomfort and suffering due to treatment complications, 5) economic, social, and psychological difficulties with the family, 6) lack of parental time, 7) lack of spiritual support, 8) influence on parental relationships (Pishkuhi et al., 2018).

Many parents express great frustration over their child’s recovery, and some think that the disease causes death and express great concern. “At first, I didn’t believe at all that my son had been diagnosed with cancer, but ever since he got worse, I’ve been constantly thinking about losing him, and I can’t sleep from these thoughts and worries at night… I have repeatedly asked God to give me death, not him” (Pishkuhi et al., 2018), Several studies have shown that chemotherapy is the worst patient experience during the treatment period. These studies show that chemotherapy, as an aggressive and severe treatment causes unwanted problems such as decreased quality of life and impaired function of parents and children. Problems of pain, hair loss, fatigue, dyspnea, and anorexia are the most problematic experiences during the treatment period from a parent’s point of view. Therefore, appropriate interventions are needed to reduce treatment complications (Pishkuhi et al., 2018).

Mothers reveal many adjustments and lifestyle changes have to live with children with leukemia. Mothers expressed concern over the fact that they had to live with a sick child with the uncertainty of the child’s prognosis. Mother expressed her feelings as “Since she was sick, I have never attended anyone’s invitations to family events and other social gatherings. I was bound by the fear that something might happen at home during my absence. That’s why I won’t go.” (Cornelio et al., 2016).

For most siblings, family is an important source of support both during treatment and pre-diagnosis. We are always there for each other. Because sometimes we really need each other. In addition, most siblings seem to feel a growing connection within the family. They have a feeling that, within their family, they can rely on one another, and that other family members are available to share their concerns, emotions and experiences (Van Schoors, De Mol, et al., 2019).

## CONCLUSION

Mothers and families play a major role in caring for and caring for children with leukemia. Must continue to take the child for check-ups to the hospital or health service. In addition, mothers also need support from their families, the environment and health workers to support and improve the level of health of their children. Mothers and families play a major role in caring for and caring for children with leukemia. Must continue to take the child for check-ups to the hospital or health service. In addition, mothers also need support from their families, the environment and health workers to support and improve the health status of their children. The study concluded that chronic diseases such as childhood leukemia have a negative impact on both the child and the mother who is the primary caregiver. Children become the focus of care in the family which can affect the welfare of other siblings in the family. The greatest burden that mothers of children with childhood leukemia face are the financial burdens associated with treatment and follow-up.

## Data Availability

All data produced in the present study are available upon reasonable request to the authors

## LIMITATION AND RECOMMENDATION

The limitation in this review is the search for qualitative study literature that discusses family experiences in caring for children with leukemia. It is hoped that this literature review can be used as literature in providing nursing interventions to families, both in the form of health education, empowerment, and support.

## ACKNOWLEDGEMENT

The authors would like to thank all the researchers whose articles were used for the discussion in this paper.

## AUTHOR CONTRIBUTION

SN participated in the analysis and further refinement of the theme; researching the literature, interpreting themes in the context of the literature and writing the first draft of the manuscript. YS participated in improving the literature review manuscript and improving the grammar used. MP participated in directing the author to write a literature review manuscript.

